# Multi-omics insights in major depressive disorder: Dysfunction of Neurons

**DOI:** 10.1101/2025.05.03.25326369

**Authors:** Lingfeng Zhang, Honggang Lyu, Simeng Ma, Yantao Xu, Xinhui Xie, Qian Gong, Lijun Kang, Shijia Chen, Zhongchun Liu

## Abstract

Major Depressive Disorder (MDD) is a complex psychiatric condition characterized by neuronal and functional disruptions in the dorsolateral prefrontal cortex (dlPFC). We integrated MDD-associated multi-omics data, including two single-cell RNA sequencing (scRNA-seq) datasets, GWAS summary data, and depression-related proteomic data from UK Biobank. We identified 273 MDD-associated eQTL, while the single-cell disease-relevance score (scDRS) algorithm revealed that excitatory neurons, inhibitory neurons, and oligodendrocyte precursor cells (OPCs) are significantly associated with MDD. Non-negative matrix factorization (NMF) identified four meta-programs (MPs) in excitatory and inhibitory neurons, reflecting functional impairments related to synaptic plasticity, neuronal connectivity, and epigenetic regulation. Differentiation trajectory analysis revealed distinct pathological states of neuronal subtypes. We identified *CXCL14^+^* inhibitory neurons and investigated their role in stress perception and intercellular communication. Plasma proteomics data validated 33 risk genes, five of which encoded proteins predictive of patient survival in CoxBoost regression models. These findings provide a comprehensive understanding of the cellular and molecular underpinnings of MDD, offering potential diagnostic and therapeutic targets and advancing the development of precision medicine approaches for MDD.

## Introduction

Major depressive disorder (MDD) is a severe psychiatric condition characterized by pervasive affective disturbances, including persistent sadness, anhedonia, and various cognitive and somatic impairments. Affecting approximately 185 million individuals globally ^1^, MDD represents a leading cause of disability, imposing a substantial burden on patients, their families, and society ^2^. Despite extensive research, the etiopathogenesis of MDD remains poorly understood and involves complex interplay interactions among genetic, environmental, and neurobiological factors ^3–5^. A significant challenge in understanding and treating MDD is the absence of robust biomarkers and the considerable heterogeneity inherent in the disorder, underscoring the need for innovative approaches to elucidate its underlying mechanisms.

MDD is a complex condition with high heritability and polygenicity ^6^. Integrating data from multiple levels is essential to minimize biases in understanding its causes. The largest depression-related GWAS to date identified 243 risk loci for depression through a meta-analysis of over 1.3 million individuals, revealing novel enrichment in neurodevelopmental and functional pathways, prenatal GABAergic neurons, astrocytes, and oligodendrocyte lineages ^7^. Whole-transcriptome and bulk RNA-seq have focused on the dorsolateral prefrontal cortex (dlPFC) and boold, revealing the role of brain and peripheral blood inflammation in depression ^8,9^. In recent years, with the advancement of transcriptomic technologies, single-cell transcriptomics and single-cell ATAC sequencing have yielded numerous findings, scaling down analyses to the cellular and epigenetic levels. These approaches have further elucidated mechanisms involved in depression, including neurogenesis, synaptic plasticity, receptor function, and neuroimmune interactions ^10–12^. However, few studies have combined multi-omics data We integrated scRNA-seq data with GWAS and eQTL findings from the dlPFC of MDD patients, validating the results via plasma proteomic data. This approach identified functional neuronal expression profiles and genes involved in neuronal development and the pathophysiology of depression. Our findings provide a comprehensive understanding of the molecular mechanisms underlying MDD and identify promising targets for future therapeutic interventions.

## Methods

### Datasets

#### GWAS datasets

We obtained GWAS summary data for mMDD from The Psychiatric Genomics Consortium (PGC), including 246,363 cases and 561,190 controls ^13^. The height GWAS data were obtained from the UKB. All participants were of European ancestry^14^.

#### eQTL Data

We obtained cis-eQTL data from multiple brain regions through the MetaBrain consortium, which provides a large-scale eQTL meta-analysis of previously published human brain eQTL datasets ^15^.

This dataset includes RNA-seq samples from 8,613 individuals, encompassing seven brain tissues (cortex, basal ganglia, hypothalamus, amygdala, hippocampus, cerebellum, and spinal cord). Significant cis-eQTLs were selected on the basis of an FDR threshold of q < 0.05.

#### scRNA-seq Data

We accessed 2 databases from the GEO database: GSE144136 and GSE 213982 and generated a gene X barcodes count matrix with 36,588 genes and 160,712 barcodes^11,12^. The study included 38 female participants (20 MDD patients (MDD), 18 health control donors (HC)) and 33 male participants (17 MDD, 16 HC) sourced from the <u>GSE144136</u> and <u>GSE213982</u> datasets, obtained from the McGill Group for Suicide Studies at McGill University ^11,12^.

#### Validation Data

This study used UKB data and Olink Explore 3072 to analyze 2,923 proteins, standardize variables and impute missing values (Supplementary File 2).

#### Mendelian Randomization

We employed Mendelian randomization to identify risk genes across multiple brain tissues, via MDD GWAS summary data and human brain eQTL datasets.

For Mendelian randomization, the Wald ratio method was applied when a single variant was available as the instrument, and the inverse-variance weighted (IVW) method with random effects was used for multiple variant instruments ^16^. The MR.RAPS robust adjusted profile score (RAPS) method was used as a confirmatory approach to enhance robustness against pleiotropic variant inclusion ^17^. Instrument selection adhered to criteria based on protein abundance or expression-associated variants with p < 5 × 10^-8^ within ±1 Mb of the coding gene, clumped at r^2^ < 0.01. To account for multiple testing, pFDR values were calculated separately for plasma protein and tissue-specific analyses. MR analyses were performed for brain gene expression (effect sizes were calculated from z scores, assuming var(y) = 1) and were deemed statistically significant at *p*FDR < 0.05. All analyses were performed using the TwoSampleMR, MendelianRandomization, and MR-RAPS packages.

### Leveraging single-cell data analysis

#### Quality control

To evaluate the quality of the barcodes, low-quality barcodes were filtered via Seurat (version 4.4.0) on the basis of mitochondrial, ribosomal, and hemoglobin gene read proportions ^18^, with the following criteria: 350 < nCount_RNA < 25,000, 200 < nFeature_RNA < 5,000, percent_ribo < 30%, and percent_mt < 5%. After filtering, 136,602 nuclei remained.

SCTransform was applied to regress out nCount_RNA, percent.ribo, percent.mt, and percent.hb, resulting in 32,186 genes and 144,531 barcodes. We calculated 100 principal components (PCs) and used the first 30 PCs for UMAP to visualize the cell populations.

Batch effects were corrected via the Harmony package (version 1.2.1) on the basis of chemical and batch information ^19^.

#### Clutsering

We optimized clustering parameters for the Seurat FindClusters function using scclusteval subsampling (80% of cells, repeated 100 times) and stability analysis via Jaccard indices. We tested k-values of 20 and 30, resolutions from 0.1 to 1.5, the smart local moving (SLM) algorithm, and Harmony-corrected PCs: 30, 40, and 50. Annotation We used specific markers to preliminarily annotate various cell types in our samples (Supplementary File 2).

#### Correct doublet effect

Initially, we identified and eliminated 7,929 high-confidence doublet barcodes via DoubletFinder (version 2.0.4) ^20^.

#### Integrate Multi-omics Data

We used the R package “scDRS” (version 1.0.2) to associate individual cells in single-cell RNA-seq data with MDD GWAS-summary data and eQTL results^13,21^. Downstream analyses utilized single-cell disease-association *P*-values and raw omics scores, to assess MDD heterogeneity within each cell type.

#### Non-negative matrix factorization for gene program discovery

The R package “GeneNMF” (version 0.6.0) was used to decompose the expression matrix of excitatory and inhibitory neurons into submatrices with k values from 2 to 6, identifying 10 MPs per dataset. Based on silhouette and Jaccard scores, MP3 and MP6 were selected for excitatory neurons, and MP4 and MP6 were selected for inhibitory neurons ^22^.

#### Pre-ranked gene set enrichment analysis

For the L4 ExN 10 and L4 ExN 12, we individually performed pre-ranked Gene Set Enrichment Analysis^23^ with clusterProfiler^24^.

### STRING analysis

We used STRING DB (version 12.0)^25^ to assess the relationships between the protein products of the overlapping genes in the DEGs, ExN MP3 and MP6.

### Trajectory Analysis

Neuronal differentiation trajectories were analyzed via “Slingshot” (version 2.10.0)^26^ and Monocle3 (version 1.3.1)^27^. SlingshotDataSet used to characterize the differentiation trajectories, and fit_models were run to identify genes that were differentially expressed by pseudotime.

### Survival analysis

Cox, random survival forest, and CoxBoost regression analyses were performed via the R packages “survival” (version 3.7-0)^28^, “randomForestRSC” (version 3.3.1)^29^, and “CoxBoost” (version 1.5)^30^ to evaluate the correlation and independence of 2,923 proteins from the UKB cohort with MDD survival time. We divided 42,712 samples (1,968 cases and 40,744 controls) into a training set (70%) and a test set (30%) and evaluated the training results using the test set. Analysis of the 33 proteins revealed five significant genes, with an AUC of 0.656 in the Cox model. Reconstructing the model with these genes improved the AUC to 0.662 in the CoxBoost model, enabling accurate prediction of the MDD survival time.

## Results

### Identification of cell types in the dPFC

We employed scRNA-seq to identify and characterize different cell types in the adult human dlPFC. Fig. 1A presents a flowchart outlining the design of our study. All the cells are illustrated and annotated with confident markers in Fig. 1B-C.

**Fig. 1.**
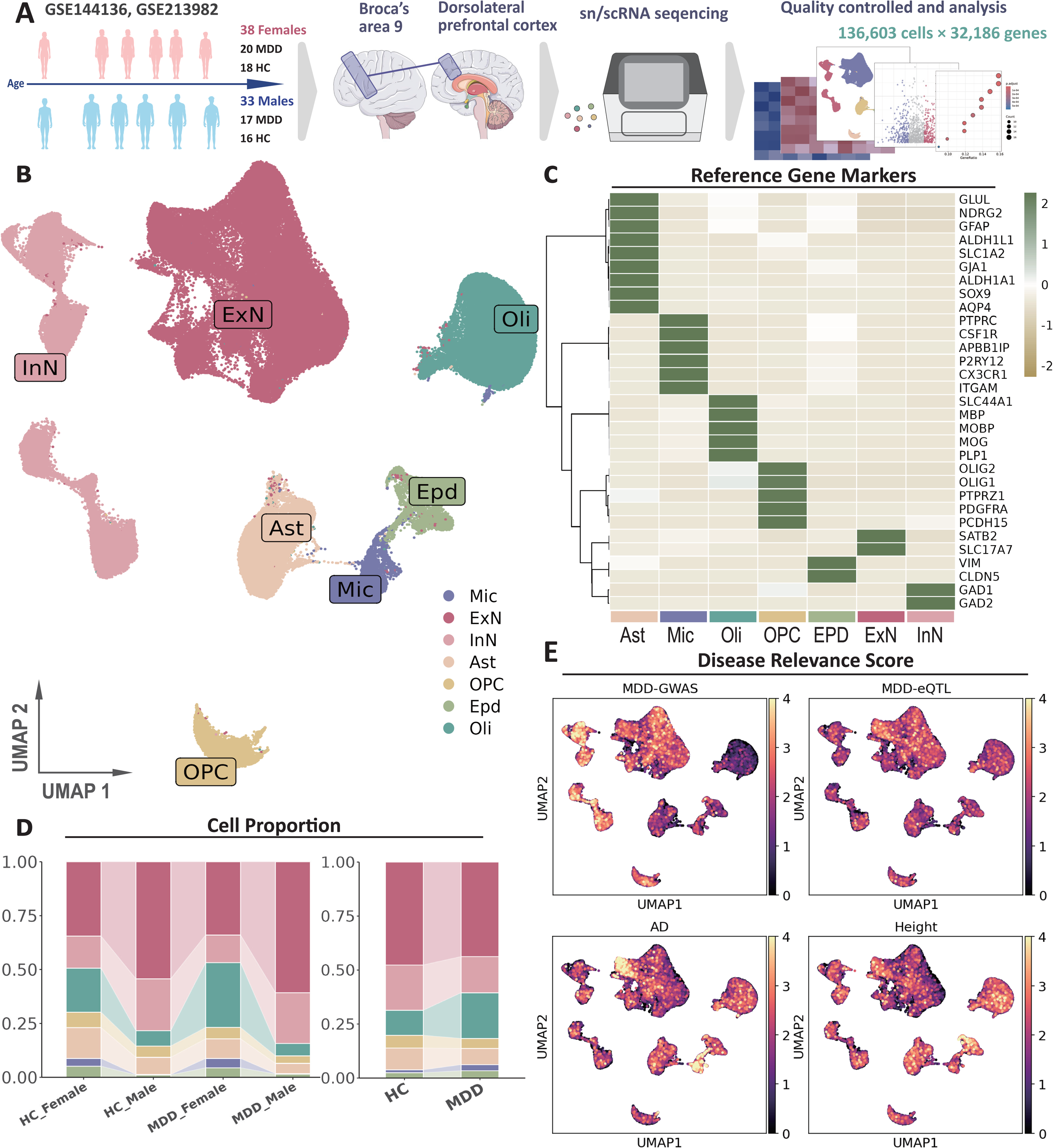
Overview and expression of MDD-related genes across different cell types in suicide MDD donors’ BA9 in the dlPFC. (A) Schematic representation of participant demographics and data sources. (B) UMAP plots depicting the identified cell types in the prefrontal cortex. (C) Heatmap of reference gene markers for each identified cell type. (D) Cell proportions by participant group (HC and MDD) and gender. (E) Disease relevance scores for the MDD-GWAS, MDD-eQTL, AD, and height groups. As a positive control, we used Alzheimer’s disease (AD) GWAS results and observed that high-risk cell subpopulations were concentrated in excitatory neurons and microglia, which is consistent with existing findings, with height-associated individuals used as controls.

The cell type proportions in the dlPFC revealed significant differences between the MDD patients and healthy controls. Specifically, the proportion of neurons in the dlPFC was significantly lower in MDD patients compared to healthy controls. This trend was observed in both male and female patients, with a more pronounced difference in female patients. Fig. 1D illustrates these differences, showing the cell proportion distributions for the healthy control and MDD groups, separated by gender. We observed that the total number of neurons in the MDD patients was significantly lower than that in the HCs.

### eQTL analysis and disease relevance Scores to identify risk cell types

On the basis of previous GWASs of MDD populations, we used cis-eQTLs as instrumental variables for Mendelian randomization (MR) analysis, identifying 273 MDD-associated genes derived from the basal ganglia, cerebellum, cortex, hippocampus, and spinal cord. We identified 150 risk genes derived from the cortex. We applied a disease-relevance score at single-cell resolution via GWAS summary and eQTL results to evaluate which cells play a critical role in the pathogenesis of MDD ^21^. Using this method, we evaluated prefrontal cortex cells and found that high-scoring cell subpopulations were predominantly found in excitatory neurons, inhibitory neurons, and the OPC, as shown in Fig. 1E.

The scDRS analysis revealed that excitatory neurons (FDR = 0.024, Z = 1.985), inhibitory neurons (FDR = 0.001, Z = 4.773), and OPC (FDR = 0.050, Z = 1.846) were positively correlated with MDD, and these correlations remained significant after multiple testing correction. To identify the most critical genes in MDD, we conducted an analysis of associated genes, and the Pearson’s correlation are listed in Supplementary File 1.

### Clustering and characterization of neurons and MPs

We applied NMF for dimensionality reduction to refine the neuronal subtypes (Fig. 2A), enhancing interpretability and ensuring that biologically meaningful metaprograms (MPs) were selected (Fig. 2B). We identified ExN MP3 and MP6 in excitatory neurons, and InN MP4 and MP6 in inhibitory neurons as key functional modules associated with MDD.

**Fig. 2.**
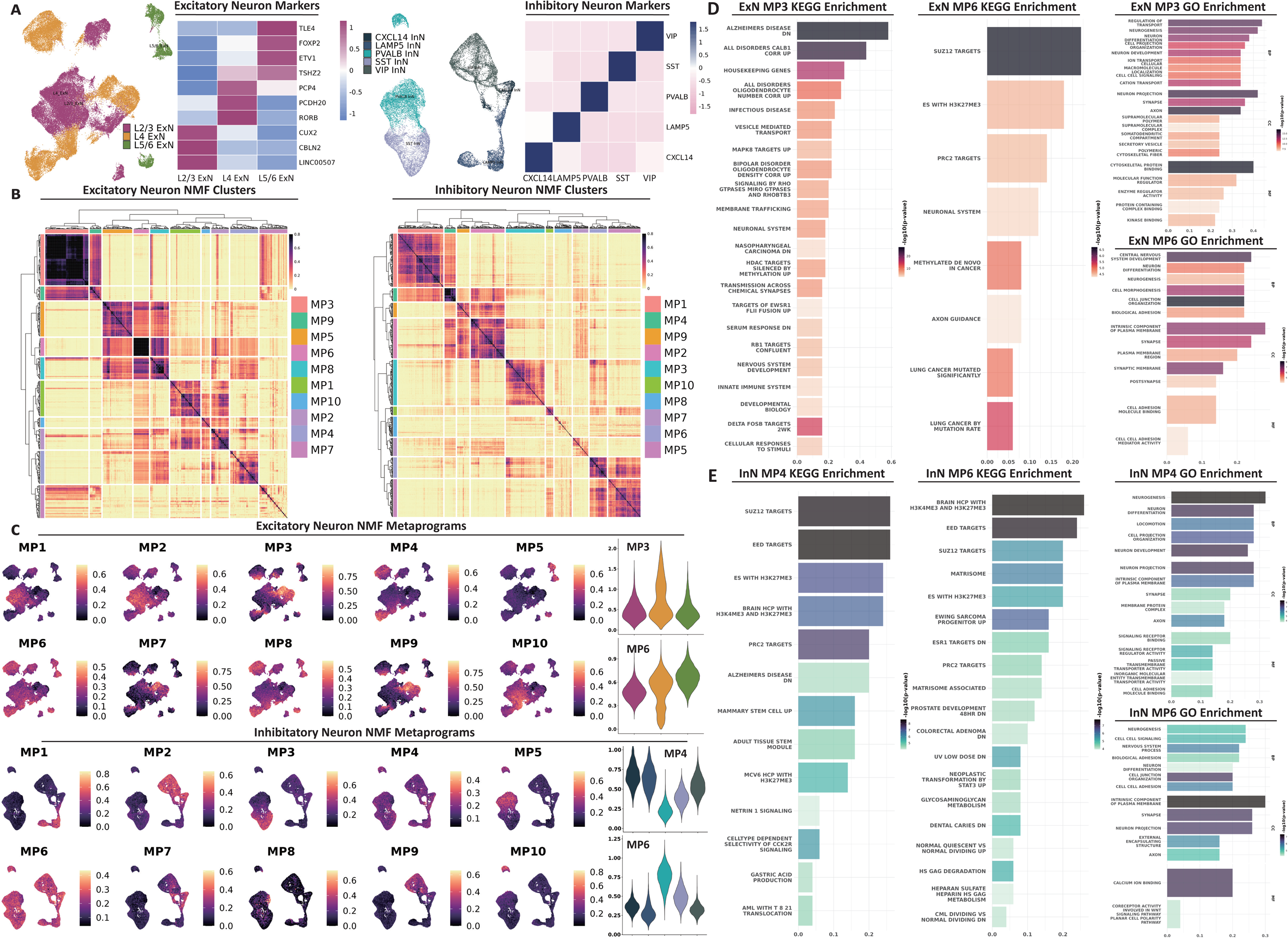
NMF embedding and identification of metaprograms in neurons of MDD patients. (A) Excitatory and inhibitory neurons, colored according to specific neuronal markers and marker gene expression. (B) NMF clustering results, showing distinct clusters of excitatory and inhibitory neuron MPs. (C) Distribution of identified metaprograms (MP1-MP10) across excitatory and inhibitory neurons. (D, E) MP functions in excitatory and inhibitory neurons.

Distinct patterns of MP expression emerged across neuronal subtypes (Fig. 2C). In excitatory neurons, MP3 and MP6 were predominantly expressed in deep-layer neurons (L4 and L5/6), suggesting their involvement in MDD pathogenesis. Among inhibitory neurons, those in the *CXCL14^+^*, *LAMP5^+^*, and *VIP^+^*subtypes primarily expressed MP4, whereas those in the *PVALB^+^* and *SST^+^* subtypes aligned more with MP6 expression, indicating distinct mechanisms through which these subtypes contribute to MDD. To investigate the role of MPs in MDD development, the enrichment analysis results are presented in Fig. 2D-E.

### Identifying the subtypes and DEGs of neurons

Building on our previous clustering analysis, we subdivided excitatory neurons into 38 subtypes and inhibitory neurons into 20 subtypes, as shown in Fig. 3A. From this refined clustering, we conducted differential gene expression analysis for each neuronal subtype, identifying 61 differentially expressed genes (DEGs) in excitatory neurons and 7 DEGs in inhibitory neurons (Fig. 3B).

**Fig. 3.**
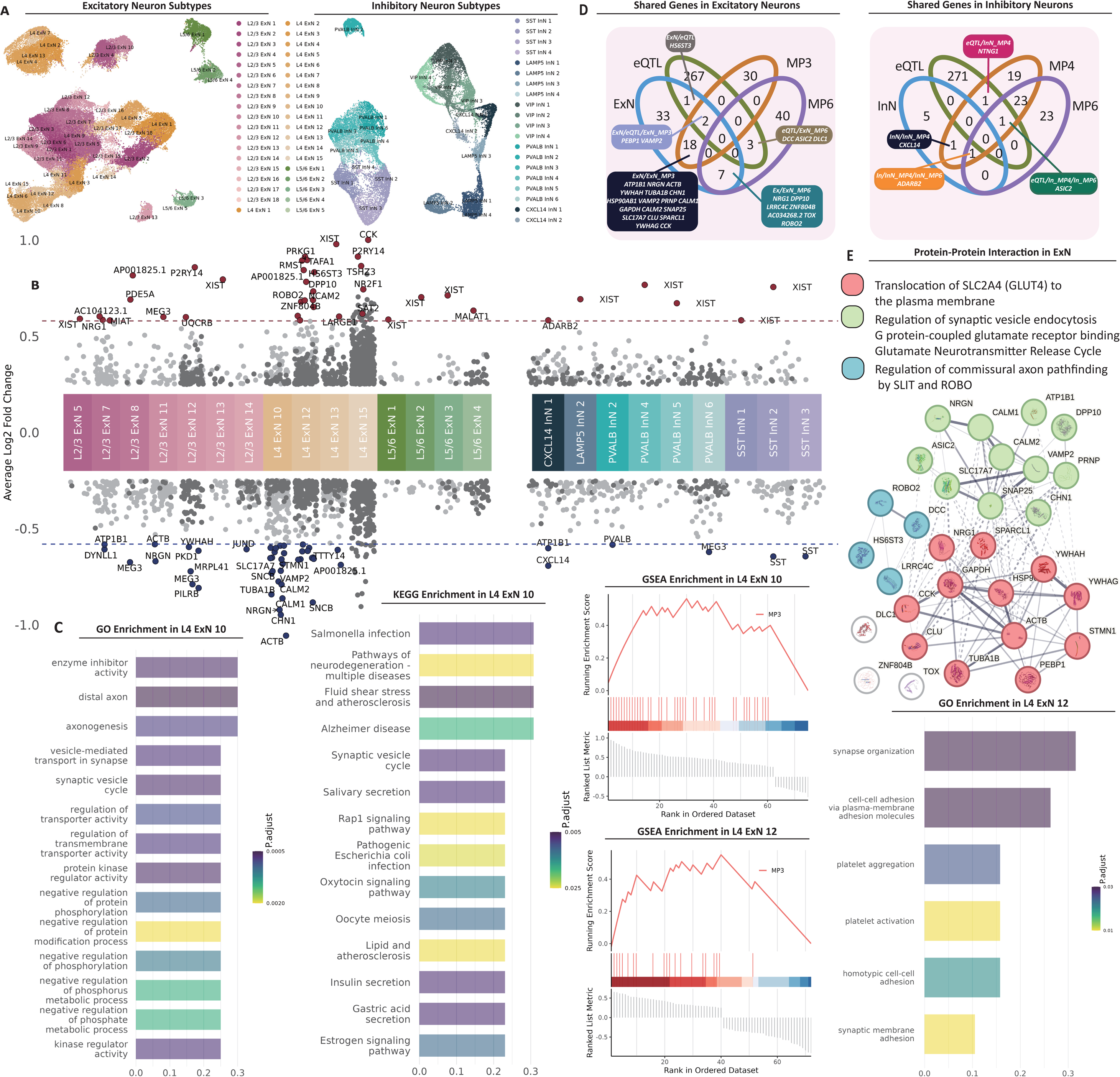
Differential gene expression and functional analysis of neuronal subtypes in MDD patients. (A) Subtypes of neurons identified in the dlPFC. (B) DEGs between MDD patients and HCs within excitatory and inhibitory neuron subtypes. (C) Functions of MDD high-risk MDD subtypes. (D) Genes that are shared across eQTLs and different metaprograms in neurons. (E) PPI network constructed from DEGs and MPs in excitatory neurons.

We identified 24 neuronal subtypes associated with MDD, including 15 excitatory and 9 inhibitory neuron subtypes. Among these, the subtypes with the most DEGs in MDD patients were L4 ExN 12, with 23 DEGs (13 upregulated and 10 downregulated), and L4 ExN 10, with 20 DEGs (all downregulated). Furthermore, we explored functions of these genes via Gene Ontology (GO), Kyoto Encyclopedia of Genes and Genomes (KEGG), and Gene Set Enrichment Analysis (GSEA) (Fig. 3C).

### Overlap genes in DEGs, eQTL Analysis and MPs

We identified significant overlaps between DEGs, eQTLs and MPs (Fig. 3D). Integration of DEGs, eQTL analysis, and ExN MP3 highlighted two genes (*PEBP1* and *STMN1*) as key molecular contributors to MDD pathophysiology. Additionally, six genes related to neuronal connectivity and epigenetic regulation consistently overlapped with the MP6 module.

In inhibitory neurons, fewer DEGs were identified. *ADARB2* and *CXCL14* are implicated in epigenetic modifications and neuronal development, whereas *ASIC2* isd linked to epigenetic regulation and cellular connectivity through overlap in the InN MP4 and MP6 modules.

### PPI clustering analysis identified key pathways associated with MDD

To understand the interactions between these overlapping genes, we conducted STRING proteinlJprotein interaction (PPI) analysis (Fig. 3E). Findings in excitatory neurons indicate the translocation of SLC2A4 (GLUT4) to the plasma membrane, the regulation of synaptic vesicle endocytosis, G protein-coupled glutamate receptor binding, the glutamate neurotransmitter release cycle, and the regulation of commissural axon pathfinding by *SLIT* and *ROBO*.

### Characterizing Neuronal Differentiation Trajectories

As shown in Fig. 4A, pseudotime analysis of neurons revealed that differentiation trajectories contributed to MDD development. While overall gene expression remained stable, some neurons transitioned from healthy to pathological states, which were absent in healthy controls. Using MDD-associated metaprograms, pathological states were labeled (Fig. 4B), and differential gene expression was visualized (Fig. 4C).

**Fig. 4.**
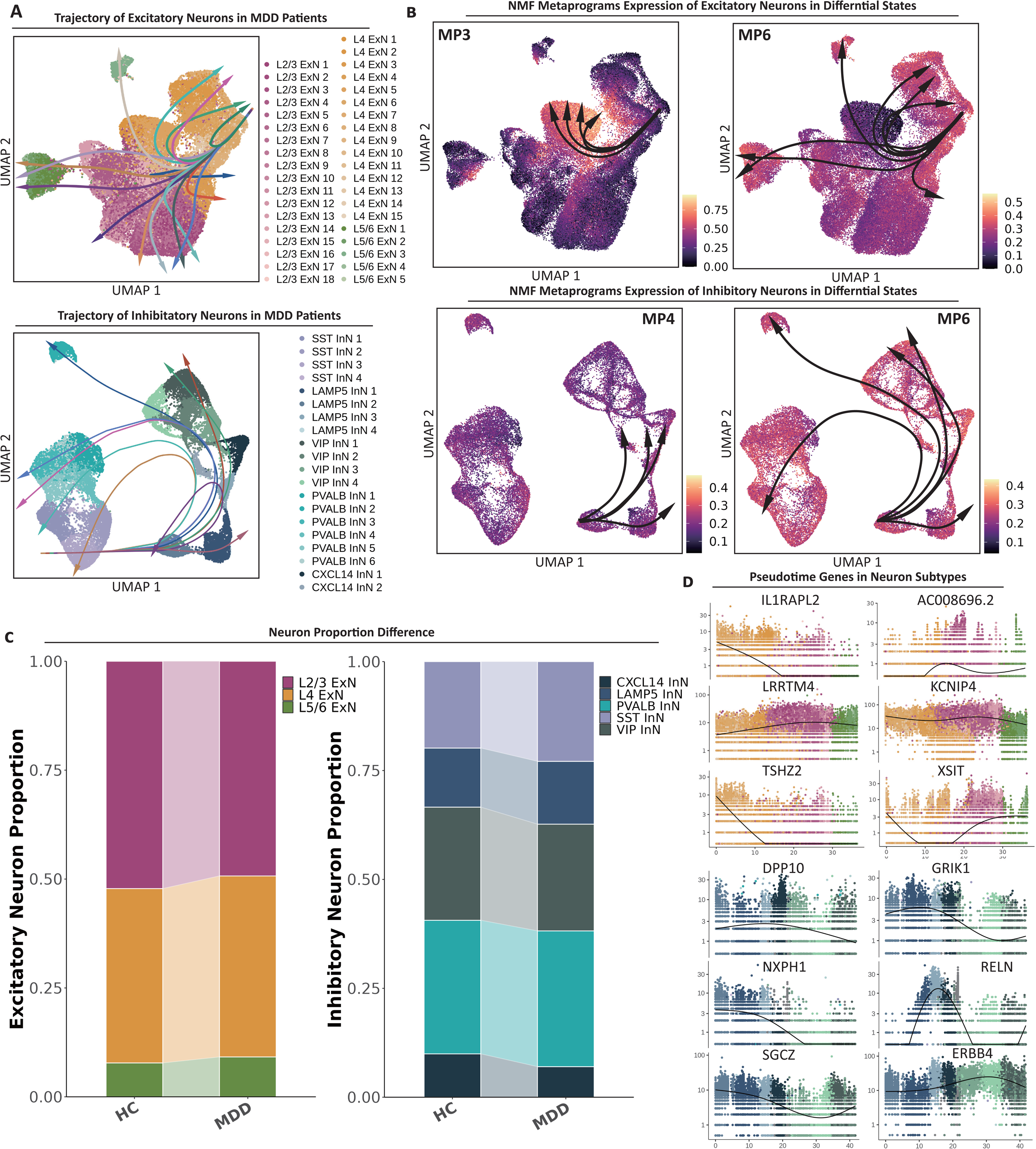
Trajectories of neuronal differentiation, metaprogram expression, and pseudotime analysis in MDD neurons. (A) Differentiation trajectories of excitatory and inhibitory neurons in MDD patients, colored by neuronal subtype, with arrows indicating progression from early to terminal states. (B) NMF metaprogram expression in excitatory and inhibitory neurons. (C) MP-associated gene expression in excitatory and inhibitory neurons. (D) Pseudotime gene expression in different subtypes.

Analysis of the top six DEGs revealed MP3 in excitatory neurons and MP4 in inhibitory neurons as intermediate pathological states, suggesting transitional dysfunction, whereas MP6 in both types was associated with terminal states (Fig. 4D). Intermediate excitatory neurons were primarily L4, and inhibitory neurons were *LAMP5^+^*and *VIP^+^*, indicating their role in early synaptic changes in MDD. Conversely, terminal states involve L5/6 excitatory neurons and *PVALB^+^* and *SST^+^* inhibitory neurons, contributing to MDD chronicity through sustained dysfunction.

### Cell Interaction Analysis and Validation in Plasma Proteomic Data

Analysis of neurotransmitter transmission (Fig. 5A) revealed increased outgoing signals from *CXCL14^+^* inhibitory neurons in MDD, with enhanced input from excitatory neurons and *LAMP5^+^* inhibitory neurons. Differentiation trajectory analysis suggested that reduced neurotransmitter transmission in deep excitatory neurons contributes to MDD onset and persistence.

**Fig. 5.**
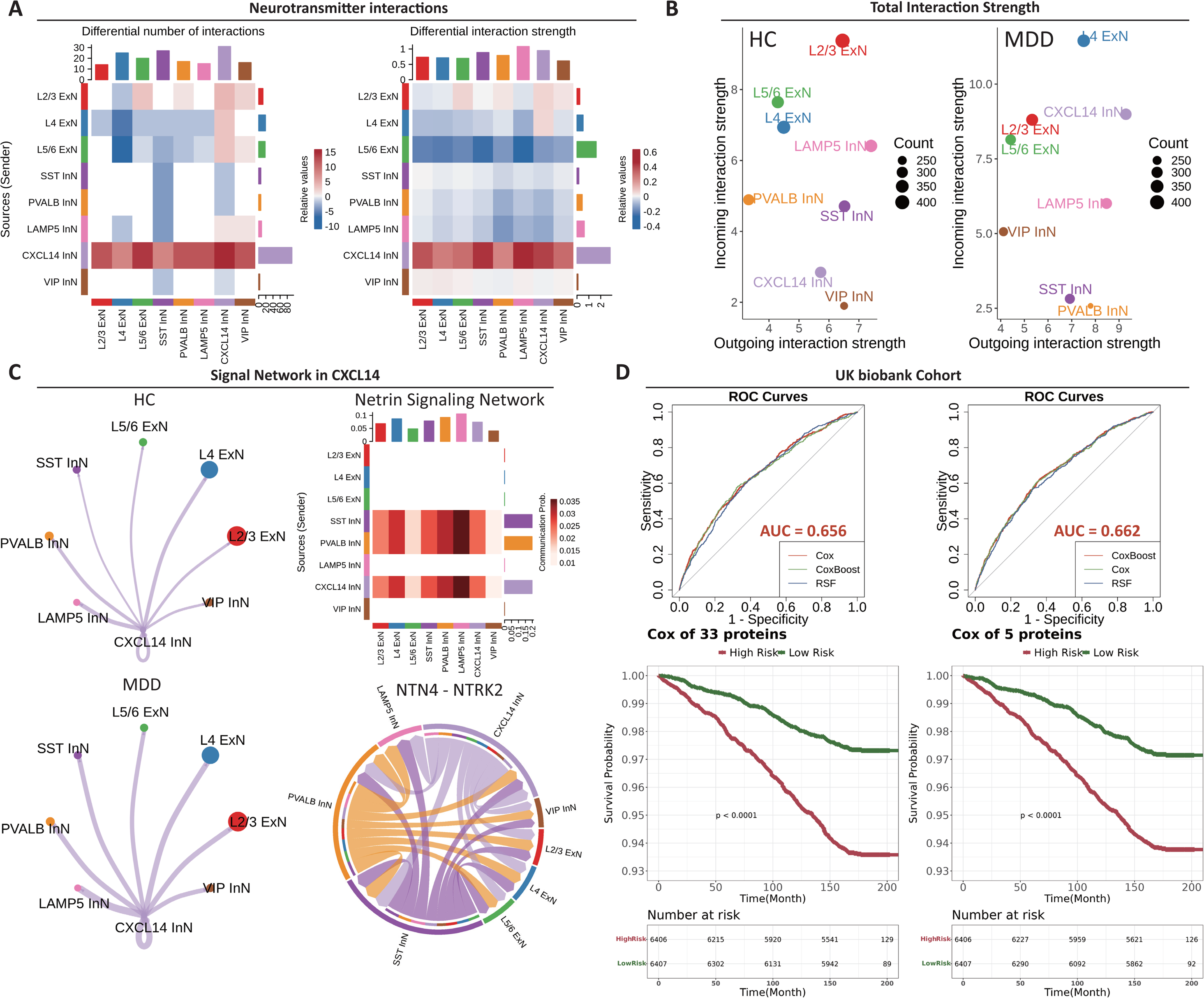
Interactions between neurons and survival analysis. (A) Neuron linked network disorders in MDD patients. (B) Differences in the signal strength of neurons. (C) The signaling network in *CXCL14^+^* neurons highlights a strong association between NTN4 and NTRK2. (D) Survival analysis of plasma proteomic data from the UKB cohort. Cox regression was performed on 33 proteins from the UKB dataset to identify 5 significant proteins, which were subsequently used for CoxBoost regression.

With respect to total neuronal interactions (Fig. 5B), L4 excitatory neurons presented increased interactions, whereas L2/3 neurons presented significant reductions, and L5/6 neurons presented decreased incoming signals. *CXCL14^+^* neurons in MDD patients also showed enhanced influence via the NTN4-NTRK2 pathway (Fig. 5C).

By integrating eQTLs, metaprograms, DEGs, and plasma proteomics, we identified 33 proteins (Fig. 5D). Regression analysis highlighted five MDD-associated proteins, with the CoxBoost model demonstrating the best prognostic performance.

## Discussion

In this study, we integrated brain region-specific eQTL data, GWAS data, and scRNA-seq data to elucidate the genetic basis of MDD. Our analysis revealed significant associations between MDD and specific neuronal populations within the dlPFC, including neurons and OPCs, at the transcriptomic level. Notably, MDD-related genes and the identified cell types presented distinct characteristics across multiple omics levels, such as differential gene expression patterns and regulatory network alterations. However, the analysis of high-dimensional scRNA-seq data poses significant challenges, particularly in identifying key biological modules. Here we employ NMF as a dimensionality reduction technique to identify biologically relevant gene expression modules and investigate their roles in MDD patients. NMF is an embedding technique that decomposes a matrix into two lower-dimensional matrices, commonly used to uncover patterns in large datasets ^31^. The raw gene counts were organized into different MPs, each representing distinct biological modules or pathways. MPs are characterized by the consistent coexpression of specific gene sets, corresponding to cellular functions, processes, or conditions, facilitating a deeper understanding of specific biological processes or disease states ^22^. Consequently, we focused on neuronal dysfunction in the dlPFC, and identified four MPs implicated in neuronal plasticity, connectivity, and epigenetic regulation. We mapped the differentiation landscape to uncover the developmental transitions involved. We revealed vital cell interactions and revealed that *CXCL14^+^*, *PVALB^+^*, and *SST^+^* inhibitory neurons in MDD patients exhibited abnormal hyperactivity in the NTN4-NTRK2 pathway. These findings underscore the pivotal role of specific neuronal subtypes in pathogenesis.

Our results indicate that both excitatory and inhibitory neurons in the dlPFC are critically involved in MDD development. The observed reduction in neuronal numbers, particularly in female patients, along with differential gene expression across neuronal subtypes, highlights the importance of targeting specific neuronal populations in pathology.

We identified a novel neuronal subtype, *CXCL14^+^* inhibitory neurons, which exhibited significant alterations in MDD patients. *CXCL14*, also known as *BRAK*, *BMAC*, or *Mip-2*_γ_, is a chemokine that belongs to the small secreted protein superfamily and plays an essential role in regulating cell migration. *CXCL14* is implicated in immune surveillance, inflammation, and tumor development ^32,33^. As a non-ELR chemokine, *CXCL14* is synthesized as a 99 amino acid precursor and subsequently processed into a mature protein with a molecular weight of 9.4 kDa ^34^. Its highly conserved sequence suggests a vital role in growth and development ^35^. *CXCL14* expression was significantly downregulated in the *CXCL14^+^* inhibitory neurons of the MDD group, indicating a marked decline in the functionality of these neurons in MDD patients. Cellular communication analysis further demonstrated that changes in *CXCL14* expression in *CXCL14^+^* neurons mirrored those observed in *PVALB^+^*and *SST^+^* neurons, which was consistent with findings from the DEG analysis. *CXCL14^+^* inhibitory neurons may be involved in modulating stress responses and maintaining neuronal homeostasis. Dysregulation of this chemokine in MDD could contribute to aberrant immune signaling and pathological processes that disrupt neuronal circuits, emphasizing its potential role in both the onset and maintenance of MDD.

To further investigate the roles of *CXCL14^+^* inhibitory neurons, we identified four MPs and characterized their respective functions. ExN MP3 represents the synaptic transmission and neuroplasticity model and is associated with neurodegenerative diseases, such as Alzheimer’s disease, suggesting overlapping mechanisms of synaptic dysfunction underlying both MDD and neurodegenerative conditions. ExN MP6 corresponds to the neuronal connectivity and epigenetic regulation model. InN MP4 and MP6 are associated with neuron projection, receptor signaling, and epigenetic dysfunctions, revealing the function of *CXCL14^+^* inhibitory neurons.

We observed strong enrichment of PRC2-related pathways in InN MP4, including targets of *SUZ12* and *EED*. These pathways regulate gene expression through chromatin modification ^36^, which is crucial for the stress response and neuronal stability. Polycomb repressive complex 2 (PRC2), a conserved epigenetic regulator of H3K27me3, plays a fundamental role in MDD pathophysiology, underscoring its potential as a therapeutic target ^36–39^. (2S,6S)-Hydroxynorketamine (HNK), the major metabolite of S-ketamine, exerts antidepressant effects through PRC2-related pathways ^37^. In rodent models, (2S,6S)-HNK modulates *SUZ12* expression, leading to antidepressant outcomes, particularly in the anterior paraventricular nucleus of the thalamus (aPVT) ^37^. These findings suggest that targeting the aPVT region may be critical for therapeutic success. The effectiveness of (2S,6S)-HNK varies across rodent models, such as the repeated tail suspension test (rTST) and chronic social defeat stress (CSDS) model, highlighting the complexity of depression mechanisms and the need for tailored treatment approaches ^37,40^.

Stress, a significant risk factor for MDD, induces long-term epigenetic changes that disrupt neuronal function by altering the expression of gens involved in synaptic plasticity. (2S,6S)-HNK counteracts these changes by restoring expression of normal gene, including *BDNF* and *CAMK2A*, to mitigate the detrimental effects of stress. Stress enhances *EZH2* activity, increasing H3K27me3 modification and stronger transcriptional repression by PRC2, which disrupts neuronal plasticity. (2S,6S)-HNK upregulates *SUZ12*, restoring PRC2 function and normalizing H3K27me3 levels ^40^.

Specifically, the observed increase in outgoing signals from *CXCL14^+^* inhibitory neurons, coupled with increased input from all excitatory neurons and *LAMP5^+^* inhibitory neurons, suggests a critical role for *CXCL14* in the disrupted signaling observed in MDD. The differentiation trajectory analysis points to reduced neurotransmitter transmission in deep excitatory neurons as a potential mechanism for the onset and persistence of MDD. Additionally, the distinct changes in interaction patterns among different layers of excitatory neurons imply that L2/3 and L5/6 excitatory neurons may be subjected to heightened inhibition from inhibitory neurons, contributing to a pathological circuit that enhances self-communication within these neurons, potentially forming a feedback loop linked to MDD.

The Netrin pathway is involved in mediating the influence of *CXCL14^+^* inhibitory neurons and other inhibitory neurons, including *PVALB^+^* and *SST^+^ neurons*. We further elucidated the significant contribution of the *NTN4-NTRK2* interaction in neurons in the MDD patients. *NTRK2*, the gene encoding TrkB, is a critical neurotrophic receptor that regulates synaptic plasticity, neuronal survival, and development by interacting with BDNF and other neurotrophic factors ^41^. Psychedelic compounds can promote significant structural plasticity in neurons by activating the TrkB, mTOR, and 5-HT2A pathways, mirroring the effects seen with rapid-acting antidepressants such as ketamine ^42^. Previous studies in the CSDS model revealed that *PVALB^+^* inhibitory neurons in the thalamus exert similar effects through the BDNFlJTrkB pathway ^43^. The stress-induced plasticity enhancement in *CXCL14^+^*, *PVALB^+^*, and *SST^+^* inhibitory neurons represents an adaptive response to stress.

However, in MDD patients, the quantity and function of *CXCL14^+^*, *PVALB^+^*, and *SST^+^* inhibitory neurons are significantly reduced. This imbalance in negative emotion regulation may be a key factor in the onset of MDD (Fig. 6.).

**Fig. 6.**
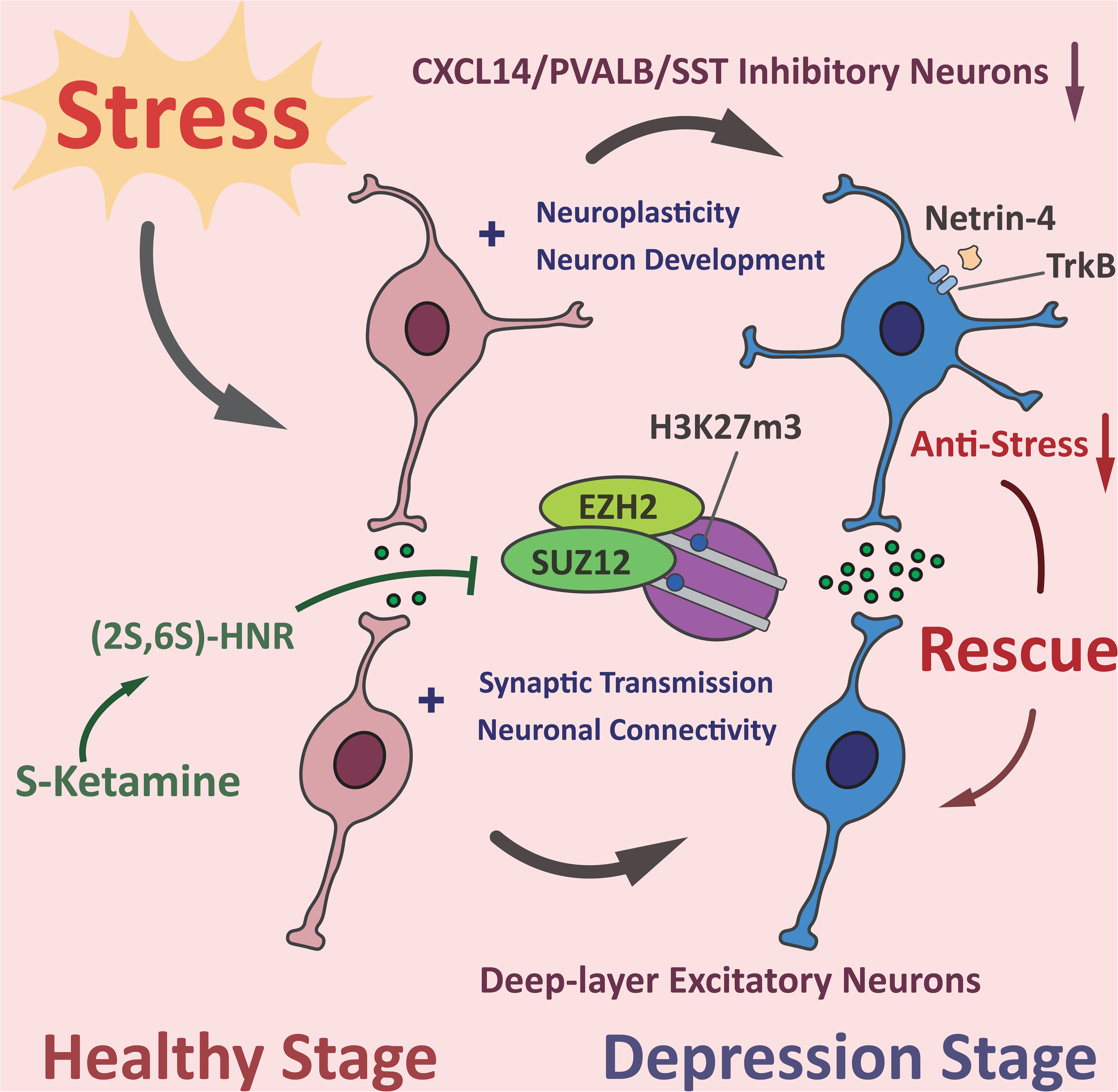
Stress-induced neuroplasticity and epigenetic regulation may play important roles in the pathophysiology of depression. Under stress, *CXCL14^+^*, *PVALB^+^*, and *SST^+^* inhibitory neurons may regulate *XIST* and the PRC2 complex, transitioning into a depressive state while enhancing synaptic plasticity in other neurons via the *NTN4-NTRK2* pathway. This pathological process is initially an adaptive mechanism to resist stress and counteract depression. However, in MDD patients, the quantity and function of *CXCL14^+^*, *PVALB^+^*, and *SST^+^*inhibitory neurons are significantly reduced, ultimately leading to the onset of depression. Antidepressants, such as S-ketamine, restore synaptic transmission and neuroplasticity, highlighting the critical roles of stress regulation, epigenetic modulation, and synaptic function in the pathogenesis and treatment of depression.

We also investigated differential neuronal subtypes and DEGs. Our findings highlight that *XIST* is highly expressed in six MDD-related excitatory neuron subtypes and five MDD-related inhibitory neuron subtypes, suggesting that *XIST* may play a significant role in the development of MDD. *XIST*, a long non-coding RNA involved in X-chromosome inactivation, recruits epigenetic regulators such as PRC2 to silence gene expression ^44–46^. While essential for dosage compensation, *XIST* also plays a broader role in the CNS, particularly in response to stress or conditions such as MDD^47^.

In L4 ExN 10, downregulated genes impaired synaptic vesicle cycling, axonogenesis, and synaptic membrane adhesion, disrupting synaptic function and neurotransmitter release. Similarly, in L4 ExN 12, downregulation affected synapse organization, celllJcell adhesion, and connectivity. GSEA revealed MP3 module enrichment in both subtypes, highlighting synaptic dysfunction and disrupted neuroplasticity as critical factors in MDD.

To validate our findings, we utilized plasma proteomic data from the UKB cohort to further investigate the functional roles of the identified risk genes. We identified 33 protein-coding risk genes in the UKB cohort and applied Cox proportional hazard models to assess their associations with survival. Our analysis revealed five proteins linked to MDD outcomes: PKD1, NEFL, and TMSB10 were associated with increased MDD risk, whereas NCAM2 and CNTN5 demonstrated protective effects against MDD. PKD1 is an integral membrane protein that functions as a calcium-permeable cation channel and a regulator of intracellular calcium homeostasis. It also plays a role in celllJcell and celllJmatrix interactions and may modulate G proteinlJcoupled signaling pathways ^48^. The NEFL protein maintains axonal stability and supports intracellular transport. It also participates in MAPK signaling cascades and modulates NMDA receptor activity, including glutamate binding and activation ^49^. NEFL is linked to neuronal damage in psychiatric conditions such as schizophrenia and bipolar disorder, indicating the occurrence of neurodegenerative processes ^50^. TMSB10 is involved in cytoskeletal dynamics and may influence neuroinflammatory pathways ^51^. In contrast, NCAM2 and CNTN5 appear to confer protective effects through their roles in promoting synaptic plasticity and enhancing neural connectivity. NCAM2 likely contributes to selective fasciculation and targeted zone-to-zone projection of primary olfactory axons ^52^.

CNTN5, an immunoglobulin superfamily member, mediates cell interactions and promotes neurite growth in cortical neurons during nervous system development ^53^.

Our study, while providing insights into MDD pathogenesis, has certain limitations. The dataset, primarily derived from Caucasian populations, limits generalizability to other ethnic groups. We used multiple datasets for intersections, which may have increased the likelihood of false negatives. Sex-specific differences in dlPFC-derived cells during MDD progression remain underexplored, despite the identification of 2 X-linked genes, *XIST* and *IL1RAPL2*. The identified MPs presented suboptimal clustering indices, reflecting the complexity of neuronal dysfunction in MDD. Furthermore, while mapping differentiation trajectories, pseudotime analysis requires experimental validation to confirm causal relationships among MPs. Owing to the cost limitations of UKB sequencing, plasma proteomics validation may be restricted to a few proteins, and neuronal-derived extracellular vesicles (NDEVs) might serve as a more specific validation method. These limitations underscore the need for further research across diverse populations and targeted investigations into the temporal dynamics of MDD pathology.

This study integrates multi-omics data to identify 273 eQTL loci, 24 differentially expressed genes in neurons, 4 MDD-associated functional modules, and 1 key pathogenic pathway, providing a comprehensive analysis of the underlying mechanisms of MDD. Validation via plasma proteomics data further highlights potential biomarkers for clinical prognosis in MDD patients.

## Declarartions

### Availability of data and materials

The MDD snRNA sequencing data can be freely accessed via the GEO database: GSE144136 and GSE 213982. Brain-region-specific eQTL results can be accessed via Supplementary File S1. The GWAS summary data can be accessed via <u>martinjzhang/scDRS</u>. Proteomics data can be accessed via UK Biobank at https://www.ukbiobank.ac.uk/.

All software used in this study is freely available and can be accessed through the provided URLs or references. The custom code developed for generating the results presented in this research is available at <u>lancelotzhang0124/SingleCell</u>.

## Data Availability

All data produced in the present study are available upon reasonable request to the authors.

## Acknowledgements

We extend our thanks to Prof. Gustavo Turecki, Dr. Corina Nagy and the McGill Group for Suicide Studies for providing the snRNA-seq data, which were crucial for our analysis. We gratefully acknowledge the support and resources provided by the Department of Psychiatry at McGill University, which were instrumental in making this research possible.

This work was supported by grants from the National Natural Science Foundation of China (U21A20364) and National Key Research and Development Project of China (Grant No. 2024YFC3308400).

---

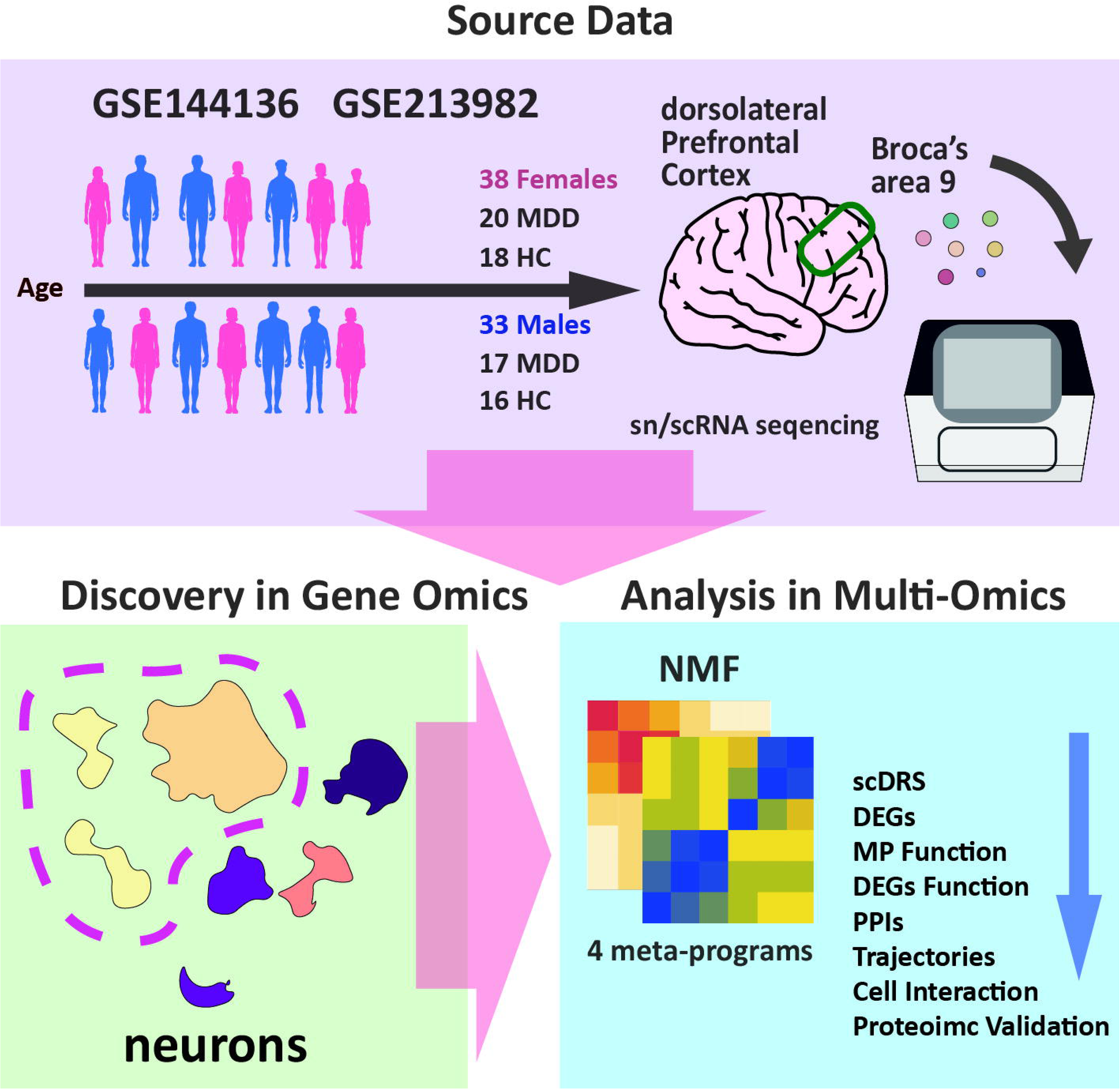

