## Supplementary File 2 for "Multi-omics insights in major depressive disorder: Dysfunction of Neurons"

**Materials and Methods**

Validation Data

Data from the UK Biobank, a large-scale biomedical resource containing genetic and health information from approximately 500,000 middle-to-older-aged British individuals, were used in this research. The UK Biobank operates under ethical approval from the North West Multicenter Research Ethics Committee (reference 11/NW/0382), with detailed research protocols available in prior publications ^1^. This analysis was conducted under UK Biobank application number 87530.

In the UK Biobank, the Olink Explore 3072 Proximity Extension Assay (PEA) platform was used to measure 2,923 protein analytes corresponding to 2,941 unique proteins across the 3,072 assays. Detailed information on these proteins is provided in Table S1. Measurements were based on Normalized Protein eXpression (NPX) values, a relative quantification unit that is logarithmically related to protein concentration, following the manufacturer's recommendations. Variables were standardized using Z-scores before analysis, and missing values were imputed with the mean.

Annotation

Excitatory neurons were identified with *SATB2* and *SLC17A7*^2,3^; inhibitory neurons with *GAD1* and *GAD2*^4,5^. Oligodendrocytes were marked by *PLP1*, *MBP*, *MOG*, and *MOBP*^6^; oligodendrocyte precursor cells by *PTPRZ1*, *PCDH15*, *OLIG1*, *OLIG2*, and *PDGFRA*^6,7^. Astrocytes were distinguished using *SLC1A2*, *ALDH1L1*, *ALDH1A1*, *GFAP*, *GLUL*, *GJA1*, *SOX9*, *AQP4*, and *NDRG2*^6,7^. Microglia were identified by *PTPRC*, *CSF1R*, *APBB1IP*, *P2RY12*, *CX3CR1*, and *ITGAM*^6,8^. Endothelial cells were characterized by *CLDN5* and *VIM*. Excitatory subtype neurons were also localized to their respective cortical layers: *CUX2*, *CBLN2*, and LINC00507 were markers for layer 2/3, *PCP4*, *PCDH20*, and *RORB* for layer 4, and *TLE4*, *FOXP2*, and *ETV1* for layer 5/6^9,10^. *TSHZ2* was identified as a marker for deep-layer neurons. Inhibitory subtype neurons were annotated based on five key markers: *CXCL14,* *LAMP5*, *PVALB*, *SST*, and *VIP*^6^.
